# Longitudinal trajectories of muscle impairments in growing boys with Duchenne muscular dystrophy

**DOI:** 10.1101/2024.06.30.24309742

**Authors:** Ines Vandekerckhove, Marleen Van den Hauwe, Tijl Dewit, Geert Molenberghs, Nathalie Goemans, Liesbeth De Waele, Anja Van Campenhout, Friedl De Groote, Kaat Desloovere

**Affiliations:** Department of Rehabilitation Sciences, KU Leuven, Leuven, Belgium; Department of Child Neurology, University Hospital Leuven, Leuven, Belgium; Clinical Motion Analysis Laboratory, University Hospital Leuven, Pellenberg, Belgium; Interuniversity Institute for Biostatistics and Statistical Bioinformatics (I-BioStat), KU Leuven, Leuven, Belgium; Interuniversity Institute for Biostatistics and Statistical Bioinformatics (I-BioStat), Data Science Institute, Hasselt University, Hasselt, Belgium; Department of Development and Regeneration, KU Leuven, Leuven, Belgium; Department of Orthopedics, University Hospital Leuven, Leuven, Belgium; Department of Movement Sciences, KU Leuven, Leuven, Belgium

**Author notes:** Corresponding author (KD).

## Abstract

**Background:** Insights into the progression of muscle impairments in growing boys with Duchenne muscular dystrophy (DMD) remains incomplete due to the frequent oversight of normal maturation as confounding factor, thereby restricting the delineation of sole pathological processes.

**Objective:** To establish longitudinal trajectories for a comprehensive integrated set of muscle impairments, including muscle weakness, contractures and muscle size alterations, whilst correcting for normal maturation, in DMD.

**Methods:** Thirty-five boys with DMD (aged 4.3-17 years) were included. Fixed dynamometry, goniometry and 3D freehand ultrasound were used to repeatedly asses lower limb muscle strength, passive range of motion (ROM) and muscle size, resulting in 165, 182 and 67 assessments for the strength, ROM and ultrasound dataset, respectively. To account for natural strength development, ROM reduction and muscle growth in growing children, muscle impairments were converted to unit-less z-scores calculated in reference to typically developing (TD) peers. This allows the interpretation of the muscle impairments as deficits or alterations with respect to TD. Mixed-effect models estimated the longitudinal change in muscle impairments.

**Results:** The pathological trajectories of most muscle impairments with age followed a similar non-linear, piecewise pattern, characterized by an initial phase of improvement or stability lasting until 6.6-9.5 years, and a subsequent decline after these ages. The muscle strength outcomes and several ROMs showed already initial deficits at young ages. General muscle weakness and plantar flexion contractures exhibited the steepest declines, resulting in large deficits at older ages. The muscle size alterations with age were muscle-specific.

**Conclusions:** The established longitudinal trajectories of muscle impairments will serve as the basis to enhance understanding of their relationship with the progressive gait pathology in DMD. Our study provides outcome measures, which will be useful for future clinical trials that assess the efficacy of novel therapeutic strategies.

## Introduction

Duchenne muscular dystrophy (DMD) is an X-linked progressive muscular disorder, affecting one per 3,500-6,000 newborn boys [1–3]. It causes dystrophin deficiency that increases muscle cell membrane fragility and vulnerability to contraction-induced damage [4–6]. Progressive muscle degeneration then emerges with loss of contractile tissue and replacement by fat and fibrotic tissue [2,7]. Boys with DMD exhibit progressive muscle impairments, including muscle size alterations, weakness and contractures [1,2,8,9]. These muscle impairments contribute to alterations in posture and gait, reflected in a motor function decline with loss of ambulation between the ages of 7.1 and 18.6 years (mean age: 12.7 years) [1,2,10]. The state-of-the-art standard of care has altered the natural disease course and has increased life expectancy in DMD [2,11–14]. One of the treatment goals in DMD is prolonging ambulation [1,2]. However, previous studies showed conflicting results on orthopedic and orthotic interventions that aim at optimizing gait and motor performance [15–19]. Since many treatment options target muscle impairments, an improved understanding of how underlying muscle impairments contribute to gait pathology in DMD is essential to improve clinical decision making and to make progress in rehabilitation, orthotic and orthopedic treatments. Furthermore, promising novel clinical trials could potentially slow down disease progression and delay loss of ambulation, but their clinical development has been hampered due to the lack of sensitive outcome measures and natural history data [20–23]. To these ends, we first need to establish longitudinal trajectories of muscle impairments. This will enrich natural history data and reveal sensitive outcome measures, and may eventually be used to enhance our understanding of their relationship with progressive gait pathology.

Progressive muscle weakness is the primary clinical symptom in DMD. McDonald et al.[24] reported that muscle strength declined at an average rate of −0.25 manual muscle testing (MMT) units over 20 muscle groups of the neck, trunk, upper and lower limb, over a period of one year, in boys aged between 5 and 13 years. However, MMT is rather subjective and appears to have restricted reliability, accuracy and sensitivity [25–27]. Objective dynamometry is, therefore, preferred for evaluating muscle weakness [28]. Significant weakness, evaluated through dynamometry, has been observed already in young boys with DMD and the strength deficit relative to typically developing (TD) children increased further with age [28–30]. This was primarily the result of the large age-related increase in muscle strength in TD, while in DMD absolute muscle strength remained relatively constant between the ages of 5 and 14 years [28,30]. After accounting for growth by normalizing muscle strength to body surface area or body mass, a significant reduction in strength with age was detected in boys with DMD [28,29]. Additionally, the use of more complex statistical models, such as non-linear piecewise models, showed that absolute muscle strength increased or remained stable until 8.5 years and then declined with age [31]. Moreover, mostly cross-sectional analyses have been performed, and therefore, caution is needed with interpreting cross-sectional age-effect as longitudinal disease progression. A previous longitudinal analysis indicated that absolute knee extension/flexion strength increased until the age of 7.5 years, while after the age of 7.5 years strength declined, with even a more pronounced decline after 9 years of age [31]. Recently, Leon et al.[32] generated percentile curves of muscle strength for boys with DMD aged 4 to 18 years based on an extended longitudinal dataset consisting of 230 assessments in 73 boys. Knee extension strength exhibited the most pronounced decline, followed by hip flexion strength, and then knee flexion strength. A recent 8-year longitudinal study of Buckon et al.[33] delineated the progressive muscle weakness in boys with DMD (aged 4 to 15 years at baseline), comparing those treated with corticosteroids (n=70) and those who were treatment-naïve (n=14). They found that knee extension strength, followed by hip extension strength, declined fastest and, that corticosteroid therapy slowed the rate of decline in knee extension strength by three times compared to treatment-naïve boys.

Apart from progressive muscle weakness, also other impairments occur in boys with DMD. Due to fat and fibrotic tissue infiltration into the muscles, prolonged static positioning and muscle weakness, boys with DMD experience progressive loss of passive range of motion (ROM), resulting in the development of contractures [8]. Whereas ankle plantar flexion contractures develop first, hip flexion, knee flexion and hip abduction contractures are also frequently present in boys with DMD [8,24,34]. In general, lower limb contractures are strongly related to wheelchair reliance [8,24,34,35]. Even though boys with DMD are longer ambulant with the widespread use of corticosteroids [36], the pattern of slow contracture evolution prior to loss of ambulation and rapid contracture evolution following loss of ambulation has persisted [24,34,35]. Choi et al.[35] found that 52.6% in the ambulatory group had ankle plantar flexion contractures and Willcocks et al.[34] showed that ROM was significantly lower in DMD than in TD, 5 years prior to loss of ambulation at the ankle, 2 years prior to loss of ambulation at the knee and 4 years prior to loss of ambulation at the hip. However, most studies focused on frequency and severity of contractures, while a quantification of the progression rate of decreasing ROM based on large longitudinal datasets is scarce. The longitudinal picture of contracture development and impact of treatment is important to be outlined, to relate these changes with the progressive gait pathology. Based on an extended longitudinal study of 1761 assessments in 322 boys with DMD, Kiefer et al.[37] found that ankle dorsiflexion ROM decreased with 1.43 degrees per year. Furthermore, Leon et al.[32] generated percentile curves for ankle dorsiflexion ROM with knee flexed and extended based on a longitudinal dataset of 258 assessments in 73 boys with DMD. The ankle dorsiflexion ROM gradually declined with age. Negative ROM values (i.e., ROM deficit to reach the neutral ankle angle of 90°) were observed from 8 years onwards with knee extended and from 9 years onwards with knee flexed, highlighting a pathological shortening in ankle plantar flexors [32]. However, previous studies did not account for the typical reduction in ROM observed in TD children aged 2 to 17 years [38–40].

The progressive loss of contractile tissue and proliferation of fat and fibrotic tissue result in varying muscle size alterations, such as profound atrophy in some muscles and (pseudo)hypertrophy in others [9]. (Pseudo)hypertrophy of the m. triceps surae is a well-known feature in DMD with reported similar contractile cross-sectional area (CSA) as in TD [9] but a total CSA up to 60% larger than in TD [28]. Yet, results on the change in m. triceps surae size with age in DMD are conflicting [9,28]. The m. quadriceps has been characterized by atrophy in DMD, with both smaller contractile and total CSA than in TD [9]. However, Mathur et al.[28] found hypertrophy of the m. quadriceps under the age of 10 years and atrophy from the age of 11 years. While the m. triceps surae and m. quadriceps represent two ends of a spectrum, the m. hamstrings and m. tibialis anterior presented with less distinctive muscle size alterations [9]. Magnetic resonance imaging (MRI) has been frequently used to quantify muscle size as well as composition and has repeatedly been suggested as sensitive biomarker in DMD clinical trials [41–45]. Ultrasound has some advantages over MRI, since it is child-friendly, relatively cheap, fast, does not require sedation, and is clinically easily accessible because it can be collected using a portable device. Ultrasound studies showed that changes in muscle echo-intensity preceded motor regression and increased over time, highlighting the added value of ultrasound as practical and child-friendly estimate of muscle composition as an outcome measure for the longitudinal follow-up of disease progression [46–48]. However, studies evaluating muscle size with ultrasound in DMD are scarce. Bulut et al.[49] reported increased muscle thickness of the m. vastus lateralis and m. medial gastrocnemius in children with DMD compared to TD children. However, Jansen et al.[47] and Pillen et al.[50] only found a reduced muscle thickness of the m. quadriceps muscle at baseline without abnormalities in thickness of the other muscles. Morse et al.[51] even showed a decrease of 36% in CSA of the m. medial gastrocnemius in adults with DMD compared to TD adults. Additionally, a longitudinal analysis indicated that muscle thickness normalized to body mass of the lower limb muscles did not progress with age [47], while another study demonstrated larger absolute muscle thickness of the m. medial gastrocnemius in more severely affected boys than in less severely affected boys [49]. This highlights that the longitudinal evolution of muscle size alterations measured by ultrasound in growing boys with DMD is still unclear.

It can be concluded that the insights into the progression of muscle impairments in boys with DMD during growth remain incomplete. This inadequacy arises from the fact that not all lower-limb muscle groups have undergone comprehensive assessment, and conflicting results among studies have been reported. Additionally, the confounding factor of normal maturation has frequently been overlooked, or various normalization techniques have been inconsistently applied. Therefore, it has been difficult to distinguish pathological changes from typical strength development, ROM reduction, and muscle growth observed in TD, hampering the interpretation of pure pathological processes during growth in DMD. We recently developed anthropometric-related TD percentile curves for muscle strength and size [52]. These curves enable us to convert individual muscle strength and size outcomes into unit- less z-scores, which indicate deficits relative to TD peers. Since this way typical strength development and normal muscle growth are accounted for, the use of z-scores facilitates the interpretation of pathological alterations during growth. Similarly, the TD reference values of Mudge et al.[38] and Sankar et al.[39] for three age groups can be used to calculate unit-less z-scores for ROM, allowing to correct for the typical reduction in ROM observed in TD. Therefore, the aim of this study was to establish longitudinal trajectories for a comprehensive integrated set of muscle impairments, including muscle weakness, contractures, and muscle size alterations, whilst correcting for normal maturation, in growing boys with DMD through a longitudinal follow-up design.

## Materials and methods

### Study design and participants

This longitudinal observational cohort study was designed with a protocol of repeated assessments with a variable number of assessments between participants and variable time intervals between assessments.

Children with DMD were recruited via the multidisciplinary clinic of the Neuromuscular Reference Centre (NMRC) in the University Hospital Leuven campus Gasthuisberg between 22 June 2018 and 31 December 2022. Boys aged between 3 and 16 years at baseline, ambulatory for at least 100 meters, and with a confirmed genetic diagnosis of DMD, were included. Cognitive and behavioral disorders preventing accurate measurements, a clinical picture of Becker muscular dystrophy, and history of muscle lengthening surgery were exclusion criteria. Use of corticosteroids and participation in clinical trials were permitted. At the University Hospital Leuven, a proactive/preventive policy was implemented to address contractures. Hereto, night-time ankle foot orthoses (AFOs) were initiated before any contractures occurred, often in conjunction with the start of corticosteroids. Occasionally, serial casting was also employed to manage early losses in ankle dorsiflexion ROM.

Retrospective data from the included children collected before 14 June 2018 (between 20 May 2015 and 14 June 2018) was accessed through the database of University Hospital Leuven between 22 June 2018 and 31 December 2022.

The local ethics committee (Ethical Committee UZ Leuven/KU Leuven; S61324) approved this study under the Declaration of Helsinki. All methods were performed in accordance with the relevant guidelines and regulations. The parents or caregivers of the participants signed a written informed consent and participants aged 12 years or older signed a written informed assent.

### Data collection and analysis

Muscle strength and size were collected unilaterally. The weakest side based on MMT was selected as the assessed side. Contractures were assessed bilaterally, but only the values of the weakest side were included in further analyses, to ensure consistency with the other measurements. If no weakest side could be identified, the assessed side was randomly chosen.

#### Anthropometry

Body mass, height and lower limb segment lengths of the boys with DMD were measured at each follow-up session.

#### Muscle weakness

A reliable and valid instrumented strength assessment was used to evaluate muscle weakness of the hip extensors, hip flexors, hip abductors, knee extensors, knee flexors, plantar flexors, and dorsiflexors [53,54]. The participants performed maximal voluntary isometric contractions (MVIC) using a fixed dynamometer (MicroFet, Hogan Health Industries, West Jordan, UT United States) in a standardized test position. Custom-written matlab (The Mathworks Inc., Natick, M.A., R2021b) scripts were used to extract the maximal force (in Newton) per MVIC and to calculate the mean maximal joint torque (in Newton meter) per muscle group by multiplying the mean maximal force over one to three representative MVIC trials with the lever arm, which was determined as 75% of the segment length. To account for typical strength development, anthropometric-related TD percentile curves for muscle strength (n=153) were used to convert mean maximal joint torques into unit-less z-scores [52]. Hence, these z-scores reflect muscle strength deficits with respect to TD peers.

#### Contractures

During a standardized clinical examination, goniometry was used to measure the passive ROM of hip extension (modified Thomas test [38]), hip adduction (with extended hip and knee of the assessed leg and hip and knee flexed in 90° of the contralateral leg), knee extension [38], hamstrings (true popliteal angle [38]), and ankle dorsiflexion (with knee extended and knee flexed in 90° [38]). Previous studies reported acceptable intra-rater and inter-rater reliability of these measures [38]. Passive ROM was measured in degrees. Since TD percentile curves for ROM are currently lacking, the age-related normative reference values of Mudge et al.[38] were used to convert ROM measures into unit-less z-scores, accounting for the typical reduction in passive ROM during growth. Thereby, the normative mean of the corresponding age group (4-7 years, 8-11 years and 12-16 years) was subtracted from the individual ROM and then this difference was divided by the standard deviation of the total normative group. We decided to use the standard deviation of the total normative group, since the large difference in standard deviations between normative age groups would result in unrealistic changes in z-scores over time. A similar approach was used to calculate z-scores for hip adduction ROM, but based on the reference values for three age groups provided by Sankar et al.[39], since these were lacking in the study of Mudge et al.[38]. By calculating the z-scores based on data for three age groups, continuous changes with age were not fully controlled for, influencing the validity of z-scores. Therefore, both absolute ROM (in degrees) and unit-less ROM (in z-scores) were reported.

#### Muscle size alterations

The reliable 3D freehand ultrasonography (3DfUS) was used to assess muscle size alterations of the m. rectus femoris, m. medial gastrocnemius and m. tibialis anterior [55,56]. The orientation and position of 2D ultrasound images were quantified using a motion tracking system (Optitrack V120:Trio, NaturalPoint Inc., Corvallis, Oregon, USA) that tracked four reflective markers on the linear probe of a Telemed-Echoblaster B-mode ultrasound device (Telemed-Echoblaster 128 Ext-1Z, with a 5.9-cm 10- MHz linear US transducer, Telemed Ltd., Vilnius, Lithuania). To limit muscle deformation, a custom-made gel pad (i.e., the portico), was attached on the ultrasound probe [57]. Data was collected and processed using STRADWIN software (version 6.0; Mechanical Engineering, Cambridge University, Cambridge, UK). The Euclidean distances between relevant anatomical landmarks were calculated to estimate muscle lengths (in mm). Mid-belly anatomical CSA (in mm^2^) was defined by one manually drawn transverse plane segmentation alongside the inner muscle border at 50% of the muscle length. We applied the same ultrasound settings and parameter definition as reported in previous studies [56,58]. In some children, high muscle degeneration strongly reduced the visibility of the muscle border. In those children, a single 2D image was collected at the mid-belly. Real-time feedback from the ultrasound images and palpation of the leg were used to identify landmarks. Subsequently, a tape measure was used to determine the midpoint between landmarks (i.e., mid-belly). To account for typical muscle growth, anthropometric-related TD percentile curves for muscle CSA (n=143) were used to convert CSA into unit-less z-scores [52]. Hence, these z-scores reflect muscle size alterations with respect to TD peers.

## Statistical analysis

To model the longitudinal change in muscle impairments, (non-)linear mixed-effect models were used.

Mixed models were fitted per outcome, i.e., strength of the 7 muscle groups (expressed as z-scores), 6 passive ROMs (absolute (in degrees) as well as unit-less (in z-scores)) and 3 muscle CSAs (expressed in z-scores). The mixed models consisted of a mean structure (i.e., fixed effects) to delineate the average trajectory and a covariance structure, induced by random effects, to describe the inter-subject variability. Age (expressed in years) at each repeated assessment represented the time-effect and was selected as the fixed effect. Random effects were included to model the variability in starting point (i.e., random intercept) and the variability in trajectory (i.e., random slope) among the subjects. We applied the previously documented workflow to construct the mixed-effect models [59–61]. Since the explorations suggested linear trends interrupted by breaking points, non-linear piecewise models were allowed. Thereby, estimated values from the explorations were used as starting values and the presence of multiple breakpoints was investigated. A breakpoint was defined if the F-test detected a significant change between adjacent slopes, whilst ensuring a minimum of 10 datapoints per regression line. Moreover, a breakpoint was not interpreted as a strict age where a change occurs, but rather as the approximate age around which a change in rate occurs. Non-nested linear and non-linear piecewise models were compared using the Akaike information criterion (AIC). Since, overall, the piecewise models demonstrated the smallest AIC, these models were selected. Yet, the difference with the linear models was small and we therefore documented the linear models in the supplementary materials (S1-S3 Figs and S1-S3 Tables). The piecewise model with observation *j* in subject *i* to estimate the outcome_ij_ was defined as follows:

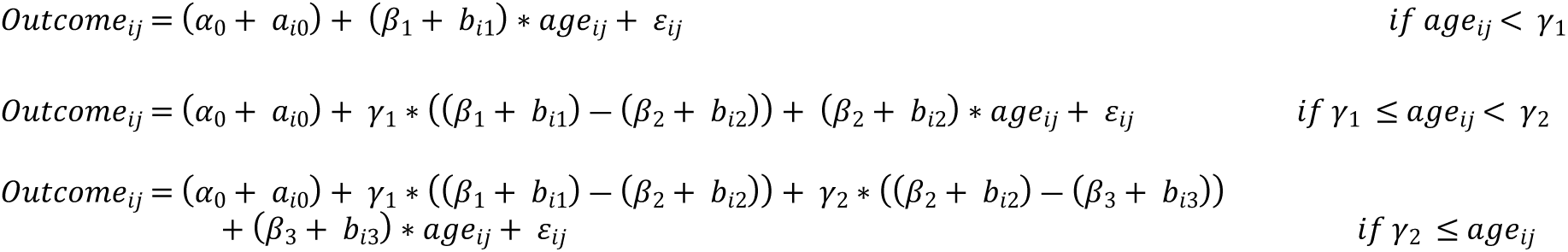

with α_0_ = intercept; a_i0_ = random intercept; β_1_ = regression slope if age_ij_ < γ_1_; b_i1_ = random slope of β1; γ_1_ = first breakpoint; β_2_ = regression slope if γ_1_ ≤ age_ij_ < γ_2_; b_i2_ = random slope of β_2_; γ_2_ = second breakpoint; β_3_ = regression slope if γ_2_ ≤ age_ij_; b_i3_ = random slope of β_3_; γ_1_ = first breakpoint; ε_ij_ = measurement error.

F-tests were performed to investigate if the intercepts, regression slopes and breakpoints differed from zero. The significance threshold was set to α = 0.05. So-called empirical Bayes estimates were calculated after model formulation and used to detect outliers, i.e., subjects with an exceptional starting point and/or evolution with age. Data of outliers were checked. SAS® (version 9.4, Statistical Analysis Software, SAS Institute Inc., Cary, NC, USA) was used to conduct all analyses and visualizations.

## Results

Thirty-five boys with DMD, aged between 4.3 and 17 years old, were repeatedly measured between 2015 and 2022 at multiple time points (1 to 11) with a variable time interval of 5-35 months, covering a follow-up period of 6 months to 6.6 years. In total, the collected dataset consisted of 185 observations (Table 1 and S4 Fig). After data collection and processing, quality check and outliers removal, the strength dataset consisted of 165 observations in 33 boys, the ROM dataset of 182 observations in 34 boys and the ultrasound dataset of 67 observations in 34 boys.

**Table 1.** Group demographics.

| Baseline characteristics |  |
| --- | --- |
| Subjects (n) | 35 |
| Median age (Q1-Q3) | 7.6 years (4.9-9.9) |
| Median body mass (Q1-Q3) | 22.4 kg (18.3-33.8) |
| Median height (Q1-Q3) | 1.16 m (1.01-1.29) |
| Median BMI (Q1-Q3) | 17.1 kg/m<sup>2</sup> (15.5-19.1) |
| Corticosteroids |  |
| Daily Deflazacort (% subjects) | 77.10% |
| Vamorolone (% subjects) | / |
| No steroids (% subjects) | 22.90% |
| Participation in clinical trial with disease modifying medication (% subjects) | 20% |
| Adherence night-time ankle foot orthoses (% subjects) | 82.90% |
| Serial casting (% subjects) | 2.90% |
| Characteristics over all observations |  |
| Subjects (n) | 35 |
| Observations (n) | 185 |
| Median age (Q1-Q3) | 10.1 years (7.6-12.5) |
| Median body mass (Q1-Q3) | 30.4 kg (22.4-40.4) |
| Median height (Q1-Q3) | 1.23 m (1.14-1.31) |
| Median BMI (Q1-Q3) | 18.9 kg/m<sup>2</sup> (16.8-24.1) |
| Corticosteroids |  |
| Daily Deflazacort (% observations) | 92.40% |
| Vamorolone (% observations) | 2.20% |
| No steroids (% observations) | 5.40% |
| Participation in clinical trial with disease modifying medication (% observations) | 36.80% |
| Adherence night-time ankle foot orthoses (% observations) | 78.90% |
| Serial casting (% observations) | 6.50% |
| Loss of ambulation (% subjects) | 31.40% |

### Muscle weakness

The longitudinal trajectories of muscle strength deficits were characterized by piecewise trends (Fig 1 and Table 2). Hip extension strength tended to increase with a rate of 0.93 z-score/year from a z-score of −3.6 z-score at 4.9 years to a z-score of −2 at age 6.6 years and then decreased with a rate of −0.29 z-score/year until a z-score of −4.9 at 16.4 years. Hip flexion strength increased with 1.03 z-score/year from −2.5 z-score at age 4.9 years to −0.7 z-score at age 6.7 years and then decreased with −0.29 z-score/year until −3.5 z-score at age 16.4 years. Hip abduction strength was −2.2 z-score at age 4.9 years and remained constant until age 8.5 years. It then decreased with −0.44 z-score/year from −1.5 z-score at age 8.5 years to −5 z-score at age 16.4 years. Knee extension strength was −0.7 z-score at age 4.6 years and remained constant until age 7.3 years. It then decreased with −0.48 z-score/year from −0.4 z-score at age 7.3 years to −4.8 z-score at age 16.4 years. Knee flexion strength was −1.5 z-score at age 4.6 years and remained constant until 8.6 years. It then decreased with a rate of −0.25 z-score/year from −1.3 z-score at age 8.6 years to −3.2 z-score at age 16.4 years. Ankle plantar flexion tended to increase with a rate of 0.25 z-score/year from −2.1 z-score at age 4.6 years to −1 z-score at age 9.1 years and then decreased with −0.39 z-score/year until −3.8 z-score at age 16.4 years. Ankle dorsiflexion strength was −2.3 z-score at age 4.6 years and remained constant until age 8.5 years. It then decreased with −0.33 z-score/year from −1.8 z-score at age 8.5 years to −4.5 z-score at age 16.4 years.

**Fig 1.**
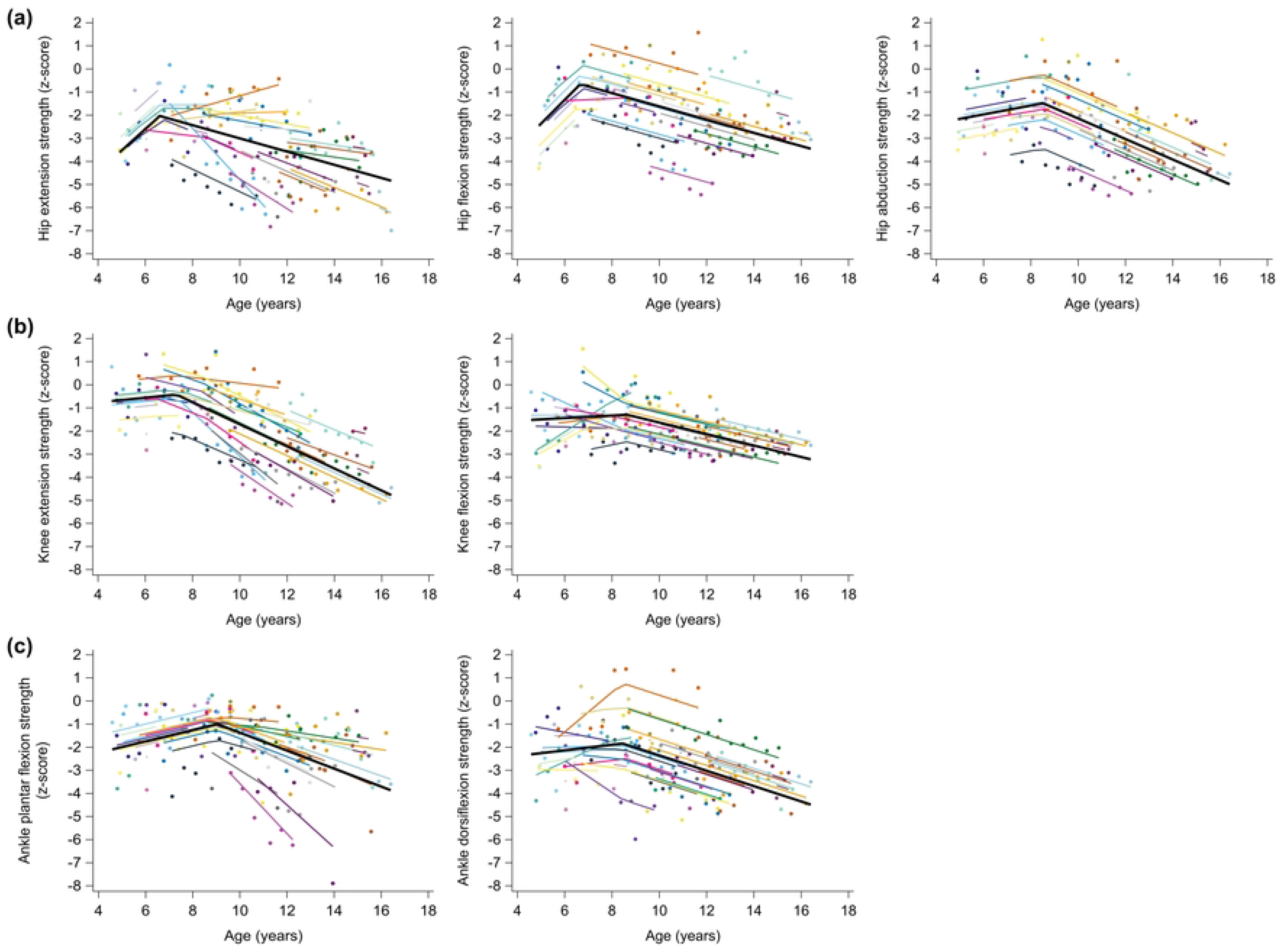
Predicted longitudinal trajectories for muscle strength deficits of hip muscles (a), knee muscles (b) and ankle muscles (c) with age for boys with DMD. The average predicted trajectory (black tick line), the individual predicted profiles (colored thinner lines), and the actual observed outcomes (colored dots) are displayed. Each color represents one patient with DMD. The estimates for the fixed effects are given in **Table 1**. DMD, Duchenne muscular dystrophy

**Table 2.** Fixed effects of piecewise models for the longitudinal trajectories of the muscle strength deficits with age for boys with DMD.

| Outcomes | n subj | n obs | Intercept | Regression coefficients ($\beta$) and breakpoints ($\gamma$) | | |
| --- | --- | --- | --- | --- | --- | --- |
| | | | $\alpha_0$ (CI)<br>*p-value* | $\beta_1$ (CI)<br>*p-value* | $\gamma_1$ (CI)<br>*p-value* | $\beta_2$ (CI)<br>*p-value* |
| Hip extension strength (z-score) | 33 | 144 | -8.16 (-13.91 -2.40)<br>**0.0070** | 0.93 (-0.05 1.90)<br>0.0613 | 6.61 (5.85 7.37)<br>**<0.0001** | -0.29 (-0.48 -0.09)<br>**0.0061** |
| Hip flexion strength (z-score) | 33 | 144 | -7.51 (-12.16 -2.87)<br>**0.0024** | 1.03 (0.23 1.83)<br>**0.0131** | 6.66 (5.90 7.41)<br>**<0.0001** | -0.29 (-0.41 -0.16)<br>**<0.0001** |
| Hip abduction strength (z-score) | 33 | 143 | -3.12 (-5.01 -1.23)<br>**0.0020** | 0.19 (-0.08 0.47)<br>0.1654 | 8.52 (7.26 9.78)<br>**<0.0001** | -0.44 (-0.57 -0.32)<br>**<0.0001** |
| Knee extension strength (z-score) | 33 | 165 | -1.18 (-2.19 -0.17)<br>**0.0234** | 0.10 (-0.05 0.26)<br>0.1813 | 7.31 (6.43 8.18)<br>**<0.0001** | -0.48 (-0.60 -0.36)<br>**<0.0001** |
| Knee flexion strength (z-score) | 33 | 165 | -1.78 (-4.16 0.61)<br>0.1389 | 0.06 (-0.23 0.34)<br>0.6953 | 8.63 (8.63 8.63)<br>**<0.0001** | -0.25 (-0.33 -0.17)<br>**<0.0001** |
| Plantar flexion strength (z-score) | 33 | 165 | -3.22 (-5.15 -1.29)<br>**0.0019** | 0.25 (-0.02 0.51)<br>0.0674 | 9.05 (7.80 10.29)<br>**<0.0001** | -0.39 (-0.58 -0.20)<br>**0.0003** |
| Dorsiflexion strength (z-score) | 33 | 165 | -2.86 (-4.90 -0.83)<br>**0.0073** | 0.12 (-0.16 0.40)<br>0.3817 | 8.45 (7.58 9.32)<br>**<0.0001** | -0.33 (-0.44 -0.23)<br>**<0.0001** |

p-values in bold indicate significance level at $p < 0.05$ .
The following symbols represent: $\alpha_0$ = intercept; $\beta_1$ = regression slope if age $< \gamma_1$ ; $\gamma_1$ = first breakpoint; $\beta_2$ = regression slope if $\gamma_1 \leq$ age.
CI, 95% confidence interval; DMD, Duchenne muscular dystrophy; n, number; o, observations; subj, subjects

### Contractures

The longitudinal trajectories of the ROMs revealed a clear degeneration solely for ankle dorsiflexion (Fig 2 and Table 3). The absolute ankle dorsiflexion ROM with knee extended was 8.3° at age 4.3 years and remained constant until 8.1 years. It then decreased with −2.7°/year from 9.7° at age 8.1 years to −8.9° at age 15.1 years, and lastly decreased with −1°/year until −10.7° at age 17 years. The unit-less ankle dorsiflexion ROM with knee extended was −2.8 z-score at age 4.3 years and remained constant until 8.6 years. It then decreased with −0.34 z-score/year from −2.6 z-score at age 8.6 years to −5.5 z-score at age 17 years. The absolute ankle dorsiflexion ROM with knee flexed was 15.7° at age 4.3 years and remained constant until age 8.1 years. It then decreased with −2.2°/year from 15° at age 8.1 years to −4.7° at age 17 years. The unit-less ankle dorsiflexion ROM with knee flexed was −2 z-score at age 4.3 years and remained constant until 8.2 years. It then decreased with −0.24 z-score/year from −2 z-score at age 8.2 years to −4.1 z-score at age 17 years. For hip extension and adduction ROM, no reliable and valid mixed models could be fitted. The data exploration (S5 Fig) suggested, however, a subtle downward trend for the absolute hip extension and adduction ROMs. At age 4.3 years, absolute and unit-less knee extension ROMs were 2.5° and −0.5 z-score, respectively. For these outcomes, no significant longitudinal change was found. Absolute hamstrings ROM was −39° at age 4.3 years, but did not progress with age. Unit-less hamstrings ROM increased with 0.10 z-score/year from −1.9 z-score at age 4.3 years to −0.6 z-score at age 17 years.

**Fig 2.**
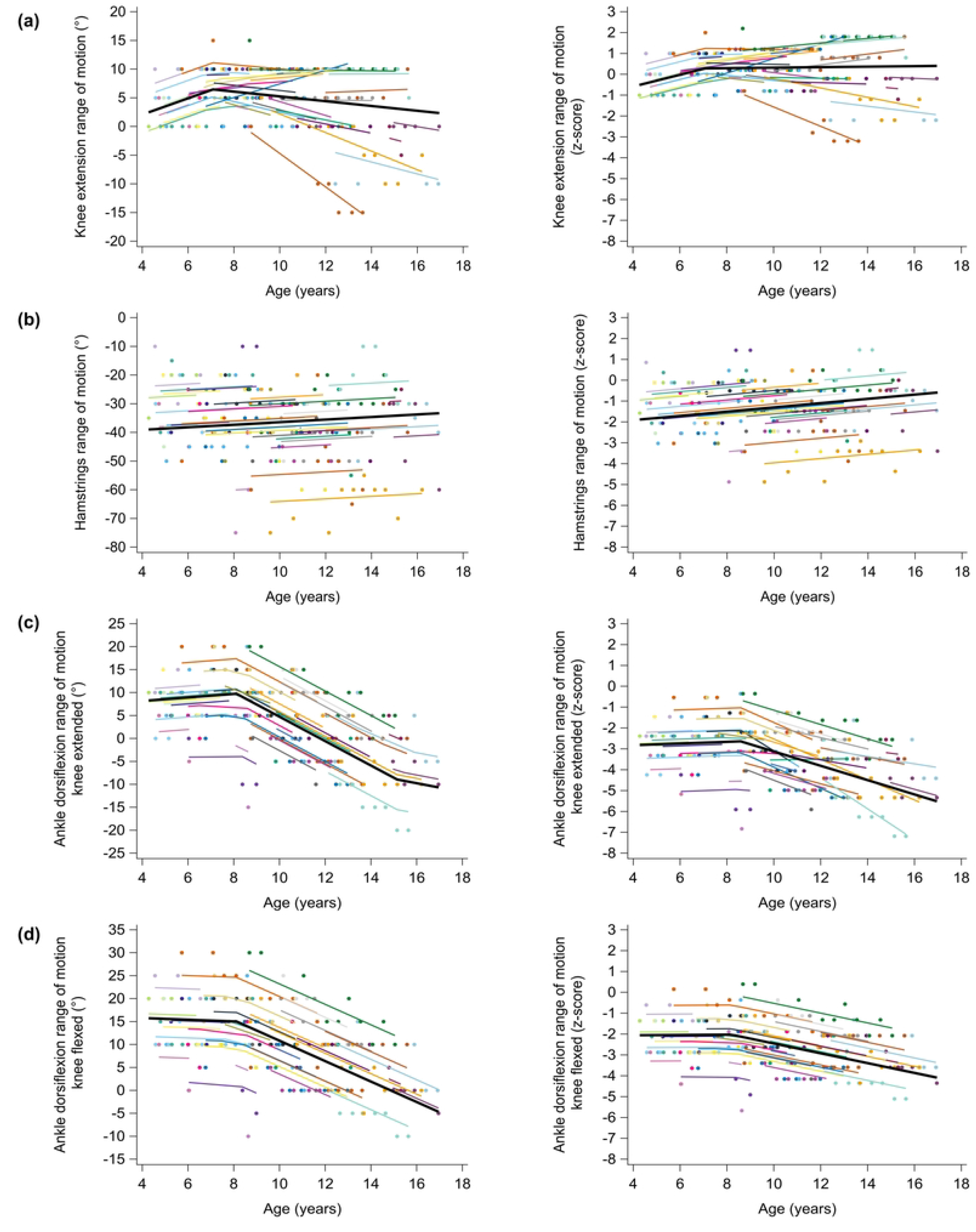
Predicted longitudinal trajectories for absolute (left column) and unit-less (right column) knee extension ROM (a), Hamstrings ROM (b), ankle dorsiflexion with knee extended (c) and ankle dorsiflexion with knee flexed (d) with age for boys with DMD. The average predicted trajectory (black tick line), the individual predicted profiles (colored thinner lines), and the actual observed outcomes (colored dots) are displayed. Each color represents one patient with DMD. The estimates for the fixed effects are given in Table 2. DMD, Duchenne muscular dystrophy

**Table 3.**
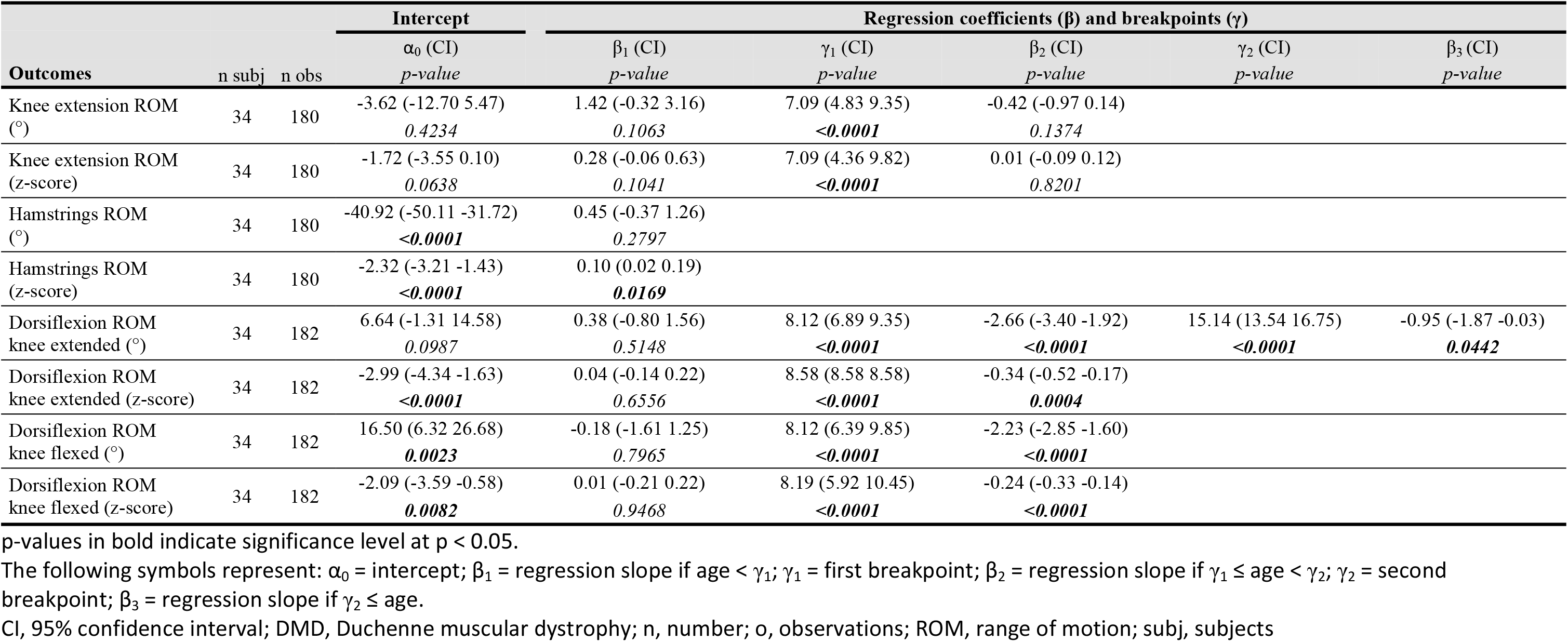
Fixed effects of piecewise models for the longitudinal trajectories of the absolute and unit-less ROMs with age for boys with DMD.

### Muscle size alterations

The longitudinal trajectories of muscle size deficits were characterized by piecewise trends (Fig 3 and Table 4). The m. rectus femoris CSA was 0.6 z-score at age 4.3 years and remained constant until age 8.8 years. It then decreased with −0.37 z-score/year from 1 z-score at age 8.8 years to −1.7 z-score at age 16.2 years. The m. medialis gastrocnemius CSA was 3 z-score at age 4.3 years and remained constant until age 9.5 years. It then decreased with −0.55 z-score/year from 4 z-score at age 9.5 years to −0.1 z-score at age 17 years. The m. tibialis anterior CSA increased with 0.46 z-score/year from −0.1 z-score at age 4.3 years to 2 z-score at age 8.8 years, then decreased with −0.45 z-score/year until 0.3 z-score at age 12.5 years, and lastly remained relatively constant until 0.3 z-score at age 17 years.

**Fig 3.**
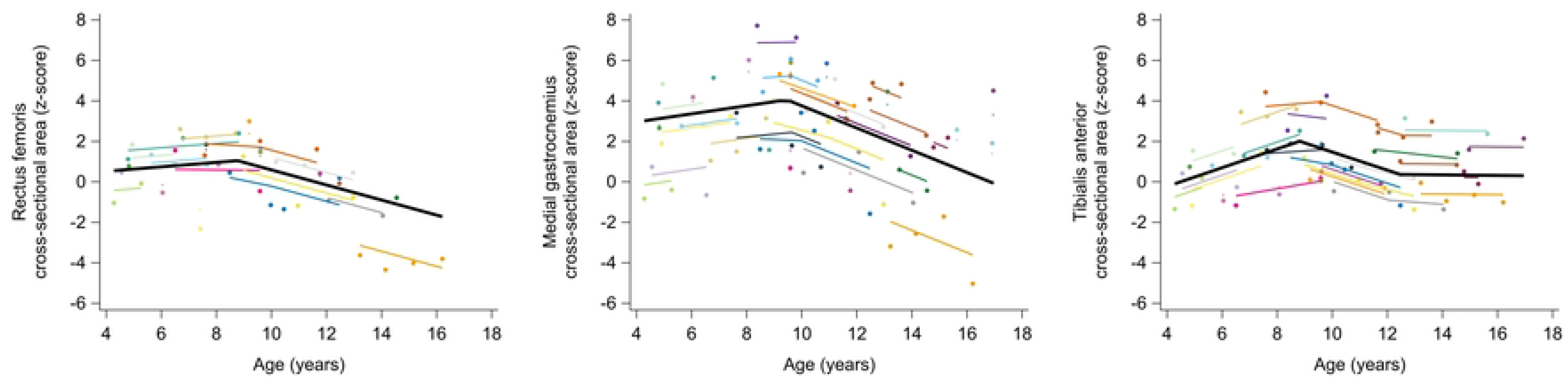
Predicted longitudinal trajectories for muscle size deficits with age for boys with DMD. The average predicted trajectory (black tick line), the individual predicted profiles (colored thinner lines), and the actual observed outcomes (colored dots) are displayed. Each color represents one patient with DMD. The estimates for the fixed effects are given in Table 3. DMD, Duchenne muscular dystrophy

**Table 4.** Fixed effects of piecewise models for the longitudinal trajectories of the muscle size alterations with age for boys with DMD.

| Outcomes | n subj | n obs | Intercept | Regression coefficients ($\beta$) and breakpoints ($\gamma$) | | | | |
| --- | --- | --- | --- | --- | --- | --- | --- | --- |
| | | | $\alpha_0$ (CI)<br>*p-value* | $\beta_1$ (CI)<br>*p-value* | $\gamma_1$ (CI)<br>*p-value* | $\beta_2$ (CI)<br>*p-value* | $\gamma_2$ (CI)<br>*p-value* | $\beta_3$ (CI)<br>*p-value* |
| Rectus femoris<br>CSA (z-score) | 24 | 46 | 0.08 (-1.75 1.91)<br>*0.9259* | 0.11 (-0.16 0.37)<br>*0.3994* | 8.77 (6.58 10.97)<br>**<0.0001** | -0.37 (-0.62 -0.12)<br>**0.0057** |  |  |
| Medial gastrocnemius<br>CSA (z-score) | 34 | 67 | 2.16 (-1.87 6.19)<br>*0.2840* | 0.20 (-0.32 0.72)<br>*0.4410* | 9.46 (7.71 11.21)<br>**<0.0001** | -0.55 (-0.89 -0.21)<br>**0.0024** |  |  |
| Tibialis anterior<br>CSA (z-score) | 29 | 64 | -2.08 (-4.64 0.48)<br>*0.1072* | 0.46 (0.11 0.81)<br>**0.0116** | 8.82 (7.77 9.87)<br>**<0.0001** | -0.45 (-0.68 -0.22)<br>**0.0004** | 12.47 (12.47 12.47)<br>**<0.0001** | -0.01 (-0.25 0.22)<br>*0.9072* |

p-values in bold indicate significance level at $p < 0.05$ .
The following symbols represent: $\alpha_0$ = intercept; $\beta_1$ = regression slope if age $< \gamma_1$ ; $\gamma_1$ = first breakpoint; $\beta_2$ = regression slope if $\gamma_1 \leq \text{age} < \gamma_2$ ; $\gamma_2$ = second breakpoint; $\beta_3$ = regression slope if $\gamma_2 \leq \text{age}$ .
CI, 95% confidence interval; CSA, cross-sectional area; DMD, Duchenne muscular dystrophy; n, number; o, observations; subj, subjects

## Discussion

This prospective longitudinal study established comprehensive trajectories of a combined set of muscle impairments, consisting of muscle strength, contractures, and muscle size alterations, in the same cohort of growing boys with DMD. We adjusted for typical strength development, ROM reduction and muscle growth observed in TD by calculating unit-less z-scores in reference to TD peers. Therefore, the established trajectories solely reflect pathological changes. This is the first study to describe longitudinal trajectories for muscle impairments expressed as deficits or alterations in reference to TD. The use of these unit-less z-scores allows for the comparison of pathological involvement between muscle groups and different muscle impairments.

Muscle weakness in growing boys with DMD follows a non-linear longitudinal trajectory, illustrating that its progression is more complex than a continuous increase. Initially, muscle strength improved (for hip flexion) or remained stable (for all other muscle groups) and then declined after 6.6-9.5 years. Lerario et al. also described muscle weakness at the knee as piecewise trends with initial improvements before 7.5 years of age and declines after. The initial improvement or stable period is consistent with the commonly held concept of the “honeymoon” period in DMD [62,63]. However, our results indicated that boys with DMD are already weaker than TD children at young ages, which aligns with previous literature [28–30]. Between muscle groups, the largest initial deficit was found for hip extension strength, followed by hip flexion, dorsiflexion, hip abduction, plantar flexion, knee flexion, and lastly knee extension strength. This only partially conforms with the known pattern of proximal to distal muscle weakness in DMD, since, unexpectedly, the ankle muscles were initially weaker than the knee muscles. After the initial improvement or stable period, the strength deficit progression rate changed to a decline at a certain age (i.e. breakpoint). Among muscle groups, this age did increase according to the proximal to distal pattern of muscle weakness (with exception of hip abduction and knee flexion). Indeed, hip extension and flexion strength began to decline at an earlier age than knee extension strength, which was followed by ankle dorsiflexion and plantar flexion strength. Yet, the decline rate was not compatible with this pattern. Specifically, knee extension strength showed the steepest decline, followed by hip abduction, plantar flexion, dorsiflexion, hip extension and flexion, and knee flexion strength. The variations in decline rate among muscle groups can be partially attributed to the timing of the breakpoint and the existing initial weakness. Previous literature reported a similar sequence of decline rate among muscle groups [32,33]. However, the longitudinal trajectories are not entirely equivalent, but due to methodological discrepancies among the studies, comparing the results proves challenging. Our results showed a flattening of the deterioration for hip extension, hip flexion and knee extension strength through the linear models (S1 Fig and S1 Table). Such a ceiling effect has also been reported by previous literature, as muscles can only degenerate until a certain point [24,47].

The trajectories of the ROMs revealed a clear decline solely for ankle dorsiflexion. In DMD, ankle plantar flexion contractures are the first contractures to develop, and, hence, occur more frequently and severely than hip and knee contractures in the ambulatory period [24,35]. Our results indicated already important plantar flexion contractures at young ages, which further progressively increased after the age of 8 years. Larger deficits were observed for dorsiflexion ROM with knee extended than with knee flexed, suggesting that the gastrocnemius is more affected than the soleus in the boys with DMD. Previous literature found that the absolute dorsiflexion ROM declined linearly from early ages onwards [32,37], which is in contrast with the non-linear piecewise trajectories in the current results. Although the observed decline rates in absolute dorsiflexion ROM were steeper than the previously reported longitudinal results of Kiefer et al. [37], the boys with DMD in the current study lost the ability to achieve neutral ankle angles at approximately 12 years of age with knee extended and at approximately 14 years of age with knee flexed, which was at an older age than reported by Leon et al.[32]. The proactive/preventive policy of addressing contractures with the early use of night-time AFOs and occasional serial casting may have played an important role in retaining dorsiflexion ROM during the initial stable phase and in delaying the inability to achieve neutral ankle angles. In contrast to the distal plantar flexion contractures, proximal lower-extremity contractures develop slowly in the ambulatory period but tend to progress rapidly after transitioning to the wheelchair [8,24,35]. The data exploration of absolute hip extension ROM and, absolute and unit-less hip adduction ROMs suggested a subtle downward trend (S5 Fig). Hip extension ROM already showed deficits in young children, indicating early hip flexion contractures, but subsequently declined at a similar rate to that observed in TD. Hip adduction ROM declined more rapidly than in TD children, yet it did not exhibit large deficits initially, implying that hip abduction contractures are absent early on but could develop after loss of ambulation. Although knee extension ROM did not progress with age and no deficit with respect to TD was found on average, there was significant inter-subject variability, and three patients presented with evident knee flexion contractures. Deficits in the hamstrings ROM were shown in young children, reflecting early hamstrings contractures, but subsequently, the ROM slightly tended to catch up that in TD children.

The trajectories of the muscle size alterations were muscle-specific, which aligns with previous literature [9]. Our results indicated that the m. rectus femoris CSA was slightly increased with respect to TD at young ages, but showed a decrease from the age of 8.8 years onwards, resulting in atrophy at older ages. This aligns with the cross-sectional findings of Mathur et al.[28], who observed hypertrophy of the m. quadriceps under the age of 10 years and atrophy from the age of 11 years. In contrast, our results indicated obvious hypertrophy in the m. medial gastrocnemius, however with a decrease from 9.5 years of age. (Pseudo)hypertrophy is a well-known symptom of DMD and has been repeatedly reported by previous studies [9,28,49]. Morse et al.[51] is, to the best of our knowledge, the only study that reported atrophy of m. medial gastrocnemius, though in adults with DMD. Hence, our observed trajectory of the m. medial gastrocnemius CSA, i.e. decreasing hypertrophy, may result in atrophy at adult ages. Although the m. rectus femoris and m. medial gastrocnemius CSAs displayed similar longitudinal trajectories, both muscles were characterized by different pathological processes (i.e., m. rectus femoris atrophy vs m. medial gastrocnemius hypertrophy), a finding also reported by the cross-sectional study of Wokke et al. [9]. In line with the observations of Wokke et al. [9], the trajectory of m. tibialis anterior CSA appeared less clearly defined with a high inter-subject variability.

This study provided novel insights by combining a set of different muscle impairments from multiple muscle groups as well as by expressing them as deficits or alterations in reference to TD, thereby solely revealing the pathological trajectories. Overall, the pathological trajectories of most muscle impairments with age followed a similar non-linear, piecewise pattern, characterized by an initial phase of improvement or stability lasting until 6.6-9.5 years, and a subsequent decline after these ages. This indicates that despite the progressive nature of DMD, the deterioration of muscle impairments is more complex than a steady decline. There is a period in which there is no deterioration (i.e., honeymoon). Both the start of corticosteroids and the preventive use of night-time AFOs may potentially play a significant role in this period of stability. However, all muscle strength outcomes (except knee extension) and several ROMs, i.e., hip extension, m. hamstrings and ankle dorsiflexion ROMs, showed already initial deficits in reference to TD early on at young ages. These proximal contractures were already present in young boys with DMD, but did not further decline with age during the ambulatory period. Conversely, general muscle weakness and plantar flexion contractures were the most important muscle impairments that exhibited the steepest deteriorations after the initial period of stability, resulting in large deficits at older ages. Moreover, the longitudinal trajectories of deficits in muscle impairments appeared to be coupled. As such, it is interesting to note the decline in muscle strength, ROM and CSA around the ankle, which all started around 8-9 years of age.

Although this study improved the insights into the longitudinal trajectories of muscle impairments in DMD based on a unique mixed longitudinal dataset, there were some limitations. The children with DMD enrolled in the study at different ages and the baseline age range was wide. Additionally, a limited number of repeated assessments was collected in some of the children with DMD. Moreover, the inter-subject heterogeneity was high due to the differences in medical and clinical background such as clinical trial participation, adherence to night-time AFOs, periods of serial casting, functional level, etc. (S4 Fig). More data is needed to delineate the influence of these background differences on the longitudinal trajectories. For the strength measurements, we were dependent on the cooperation and motivation of the children. Caution is needed with the interpretation of the unit-less ROMs trajectories, as they were calculated based on age categories, which could result in abrupt changes in the unit-less ROMs with age. This highlights the need for TD percentile curves for ROMs to aid in distinguishing pathological reductions in ROM from the typical reduction in ROM during growth. Lastly, the sample size of the ultrasound dataset was small, especially for the m. rectus femoris, since we were unable to process the data of some older patients with DMD due to muscle border invisibility caused by severe muscle degeneration. Therefore, the results of ultrasound dataset should be interpreted with caution.

Despite the limitations, this study has established longitudinal trajectories of muscle impairments, consisting of muscle strength, contractures and muscle size alterations, in a cohort of growing boys with DMD covering the ambulation period. These longitudinal trajectories are valuable for enhancing our understanding of the relationship between muscle impairments and progressive gait pathology in DMD. Moreover, the results revealed sensitive outcome measures, which could enhance clinical trial design to detect the efficacy of novel therapeutic strategies.

## Data Availability

The data supporting the findings of this study are openly available in the Research Data Repository (RDR) of KU Leuven at https://doi.org/10.48804/ONRNG0.

## Acknowledgments

The authors wish to thank all the children and parents for their participation in this study. We also thank the colleagues of the University Hospital of Leuven involved in the recruitment. A special thank you to Elze Stoop and Ester Huyghe for their support in data collection and processing.

## Supporting information

**S1 Fig. Predicted longitudinal trajectories for muscle strength deficits of hip muscles (a), knee muscles (b) and ankle muscles (c) with age for boys with DMD of linear mixed effect models.** The average predicted trajectory (black thick line), the individual predicted profiles (colored thinner lines), and the actual observed outcomes (colored dots) are displayed. Each color represents one patient with DMD. The estimates for the fixed effects are given in Supplementary Table 1. DMD, Duchenne muscular dystrophy

**S1 Table. Fixed effects of linear mixed effect models for the longitudinal trajectories of the muscle strength deficits with age for boys with DMD**

p-values in bold indicate significance level at p < 0.05.

The following symbols represent: α_0_ = intercept; β_1_ = regression slope of age; β_2_ = regression slope of age^2^; β_3_ = regression slope of age^3^. CI, 95% confidence interval; DMD, Duchenne muscular dystrophy; n, number; o, observations; subj, subjects

**S2 Fig. Predicted longitudinal trajectories for absolute (left column) and non-dimensional (right column) knee extension ROM (a), Hamstrings ROM (b), ankle dorsiflexion with knee extended (c) and ankle dorsiflexion with knee flexed (d) with age for boys with DMD of linear mixed effect models.** The average predicted trajectory (black thick line), the individual predicted profiles (colored thinner lines), and the actual observed outcomes (colored dots) are displayed. Each color represents one patient with DMD. The estimates for the fixed effects are given in Supplementary Table 2. DMD, Duchenne muscular dystrophy; ROM, range of motion.

**S2 Table. Fixed effects of linear mixed effect models for the longitudinal trajectories of the absolute and non-dimensional ROMs with age for boys with DMD**

p-values in bold indicate significance level at p < 0.05.

The following symbols represent: α_0_ = intercept; β_1_ = regression slope of age; β_2_ = regression slope of age^2^; β_3_ = regression slope of age^3^.

CI, 95% confidence interval; DMD, Duchenne muscular dystrophy; n, number; o, observations; ROM, range of motion; subj, subjects

**S3 Fig. Predicted longitudinal trajectories for muscle size deficits with age for boys with DMD of linear mixed effect models.** The average predicted trajectory (black thick line), the individual predicted profiles (colored thinner lines), and the actual observed outcomes (colored dots) are displayed. Each color represents one patient with DMD. The estimates for the fixed effects are given in Supplementary Table 3. DMD, Duchenne muscular dystrophy

**S3 Table. Fixed effects of linear mixed-effect models for the longitudinal trajectories of the muscle size alterations with age for boys with DMD**

p-values in bold indicate significance level at p < 0.05.

The following symbols represent: α_0_ = intercept; β_1_ = regression slope of age; β_2_ = regression slope of age^2^.

CI, 95% confidence interval; CSA, cross-sectional area; DMD, Duchenne muscular dystrophy; n, number; o, observations; subj, subjects

**S4 Fig. Overview of the participants medical and clinical background. Each color represents one patient with DMD.**

AFO, ankle foot orthosis; DMD, Duchenne muscular dystrophy; LA, loss of ambulation

**S5 Fig. Data exploration of longitudinal trajectories for absolute (left column) and non-dimensional (right column) hip extension ROM (a) and hip adduction ROM (b) with age for boys with DMD of linear mixed effect models.** The Loess regression (black thick line) and the actual observed outcomes (colored symbols) are displayed. Each color represents one patient with DMD. DMD, Duchenne muscular dystrophy; ROM, range of motion.

**S4 Table. Estimates of random-effect and residual covariance structure of piecewise models for the longitudinal trajectories of the muscle impairments with age for boys with DMD**

The following symbols represent: σ^2^ =variance; a_i0_ = random intercept; b_i_ = random slope; b_i1_ = random slope for regression slope before breakpoint; b_i2_ = random slope for regression slope after breakpoint; ε_ij_ = measurement error.

CSA, cross-sectional area; DMD, Duchenne muscular dystrophy; ROM, range of motion;

**S5 Table. Estimates of random-effect and residual covariance structure of linear mixed-effect models for the longitudinal trajectories of the muscle impairments with age for boys with DMD**

The following symbols represent: σ^2^ =variance; a_i0_ = random intercept; b_i1_ = random slope for regression slope of age; ε_ij_ = measurement error.

CSA, cross-sectional area; DMD, Duchenne muscular dystrophy; ROM, range of motion;

